# Not Forgotten: Patient Experiences with Genetic Variant Reclassifications

**DOI:** 10.64898/2026.05.06.26352483

**Authors:** Pankhuri Gupta, Min Seon Park, Eric Y. Kao, Abbye E. McEwen, Runjun D. Kumar, Martha Horike-Pyne, Douglas M. Fowler, Lea M. Starita, Sarah Knerr, Andrew B. Stergachis

## Abstract

**Purpose:** Genetic variant reclassification is increasingly common in clinical genomics, yet limited data describe how patients experience re-contact and variant reclassification in routine clinical care.

**Methods:** We conducted semi-structured qualitative interviews with 20 adult patients who received a variant reclassification following routine clinical genetic testing. Interviews explored emotional responses, communication experiences, and perceived value of genetic testing. Data were analyzed using Template Analysis, a form of thematic analysis.

**Results:** Three overarching themes were identified. Participants identified a need for improved communication of reclassified results, particularly with respect to timing, modality, and contextualization (Theme 1). Experiences with reclassification also shaped perceptions of the value of genetic testing, with most participants viewing testing as worthwhile despite its evolving nature (Theme 2). Finally, many participants interpreted reclassification as evidence of personalized and ongoing care, reinforcing trust in genetic testing and biomedical research (Theme 3). Participants generally preferred to be informed of reclassified results regardless of reclassification type, although the direction of reclassification influenced emotional responses and preferred modes of communication. Downgrades from variants of uncertain significance to benign or likely benign were widely viewed as meaningful by participants.

**Conclusion:** Variant reclassification was experienced as a signal of personalized, ongoing care. Timely, contextualized, patient-centered re-contact practices may reduce uncertainty, strengthen trust, and help patients not feel forgotten.

## Introduction

Updates to variant classification criteria, together with advances in genomics—including more inclusive population databases, multiplexed functional assays, and improved computational predictors—have enabled the refinement of genetic variant classifications over time^1–5^. These refinements are foundational to personalized genomic medicine and have extended its benefits to a broader range of individuals. However, a consequence of this progress is that the classification of a genetic variant reported to a patient may change as new evidence accumulates. As a result, variants initially reported as a variant of uncertain significance (VUS) may later be resolved as being pathogenic or benign, or previously reported pathogenic or benign variants may be downgraded or upgraded, respectively^6^.

Variant reclassification is an increasingly common occurrence in clinical genomics. Recent studies estimate that approximately 4.7% of all VUS in ClinVar have been reclassified since 2020^7^, with 3-4.5% of VUS undergoing reclassification in each two-year period^8^. Despite the integral role of variant reclassification in genomic medicine, clinical practices for returning reclassified results remain inconsistent^9^. While the 2024 ACMG “Points to Consider” statement frames patient re-contact as a shared responsibility among the provider, laboratory, and patient^10^, implementation in practice is variable. As a result, reclassifications affecting up to 1.6% of individuals in one health system were never communicated to the patient or their provider^11^.

Beyond logistical challenges, variant reclassification raises important clinical and psychosocial questions. Prior work has demonstrated that reclassification can alter clinical management recommendations^12–14^. In addition, reclassification may carry emotional and behavioral implications for patients. A growing body of research has begun to examine patient perspectives on these experiences. For example, interviews with patients and family members enrolled in a VUS reclassification research study found that participants were often motivated by a desire for answers and generally responded positively when a VUS was reclassified to a more definitive result^15^. Similarly, another study found that most participants did not experience distress following variant reclassification in hereditary cancer genes, and only a minority reported decreased trust in medical genetics^16^. Nonetheless, negative psychological outcomes were reported in cases where reclassification was inconsistent with patients’ prior expectations^17^. In addition, a survey of patients with VUS classifications found that participants largely indicated a preference for written disclosure for VUS downgrades but a combination of written and verbal disclosure for VUS upgrades^18^.

Despite these insights, there remains limited in-depth understanding of how patients interpret and emotionally process reclassified results outside of research settings and across different reclassification trajectories and clinical contexts. Understanding how patients perceive dynamic genetic test results is essential for genetics providers and healthcare systems navigating recontact practices. To address these gaps, we conducted a qualitative study exploring how patients experience and respond to reclassification of their genetic test results. Specifically, we examined: (1) the effectiveness of communication surrounding variant reclassification; (2) participants’ trust in the healthcare system in the context of reclassification; and (3) the perceived value of genetic testing following reclassification. Our findings inform policy and clinical recommendations to improve re-contact processes and better support patients within the rapidly evolving landscape of genomic medicine.

## Methods

This study was approved by the Institutional Review Board (IRB) at the University of Washington (STUDY00022283). Eligible participants were English-speaking adults (18 years or older) who had undergone clinical genetic testing at the University of Washington (UW) or Fred Hutchinson Cancer Center (FHCC) and had been informed of a variant reclassification within the prior two years.

To identify potential participants, a list of patients who had received a variant reclassification in the past two years from a germline panel-based genetic testing was requested from an external commercial genetics laboratory. Chart review was performed to confirm study eligibility. Disclosure of reclassified results was verified through EHR documentation by the clinician, either as a note shared directly with the participant or as a progress note describing an inperson or telehealth visit where the result was shared. Indication for testing (*i.e*., personal history [PH] or family history [FH] or both) was also captured via EHR documentation. Evidence for reclassification (e.g., new functional data, updated population databases, additional family/trio sequencing data) was not captured since such information was not included as a part of the report that the laboratory issued.

Eligible individuals were contacted by a genetic counselor via email or phone to invite them to participate in the study up to three times. Recruitment was performed on a rolling basis until our target sample size of 20 participants was achieved (**Fig. 1A**). Convenience sampling was used and the first 20 individuals who consented were included in the study.

**Figure 1.**
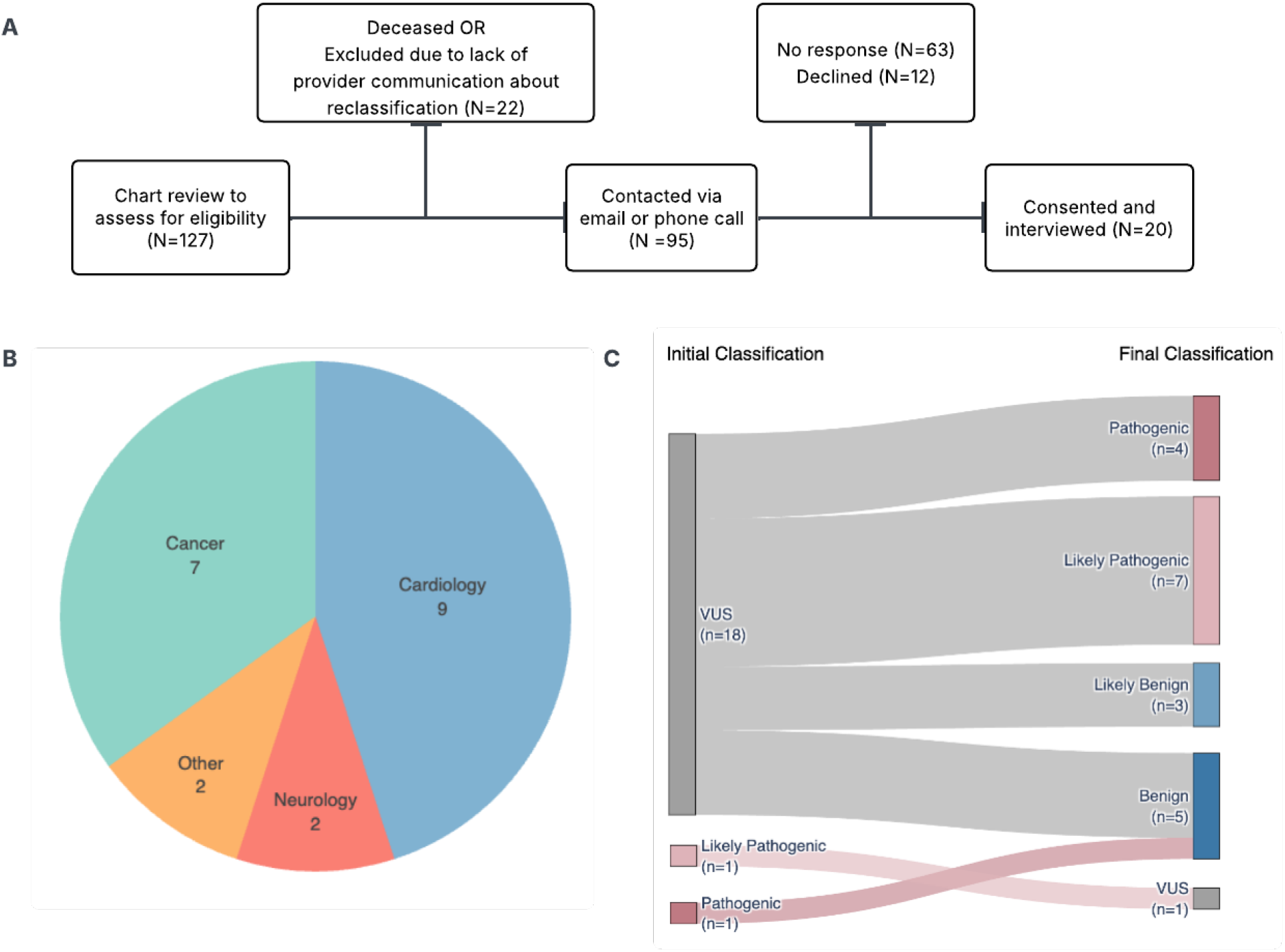
Study cohort characteristics. (A) Flow diagram of participant identification, eligibility assessment, and enrollment. (B) Distribution of clinical indications for participants. (C) Sankey diagram showing changes between initial and final variant classifications.

### Interview and Data Collection

We developed a semi-structured interview guide (**Supplemental Document**) focused on four key domains: (1) emotional reactions to the reclassified result, (2) understanding and interpretation of the new information, (3) support and resources provided, and (4) reflections on the overall value of genetic testing. While the interview guide was followed to ensure that all relevant questions were asked, further probing questions were also included to ensure detailed narratives.

Interviews were conducted via Zoom or telephone by a genetic counselor (GC) who was not part of the participant’s clinical care team between May and August 2025 and were audiorecorded with verbal consent. Zoom-generated transcripts were reviewed for accuracy against the recordings.

### Data Analysis

We conducted a Template Analysis^19^, a form of thematic analysis, following the iterative steps outlined by King^20^. First, two GC investigators independently read the transcripts and noted initial impressions that were related to the research questions: (1) the effectiveness of communication surrounding variant reclassification; (2) participants’ trust in the healthcare system in the context of reclassification; and (3) the perceived value of genetic testing following reclassification. Next, preliminary open coding was performed by assigning codes to meaningful units of text (such as phrases indicating an emotion or an action), which were organized and used to define an initial coding template. The initial coding template was updated iteratively until both GC investigators agreed with the codes identified. Throughout coding, the GC investigators regularly discussed code definitions and resolved discrepancies, ensuring consistent application of codes across transcripts. Emotional responses were coded using participants’ own words where possible, or mapped to the closest existing code in the developed codebook. The frequency of each emotion was then compared across the two reclassification groups within those who had an initial VUS result. Ideas that could not be captured in distinct codes were noted as unique memos. Two GC investigators coded 7 transcripts together, with the remaining 13 transcripts divided between the two investigators for independent coding.

Once all transcripts had been coded, codes and memos that were conceptually related were grouped together into themes and iteratively refined. Each theme was defined to ensure that it reflected a distinct aspect of the overall data. The qualitative data management software Dedoose (v. 10.0.59) was used to facilitate coding and thematic analysis.

## Results

### Clinical characteristics of participants and their variant reclassifications

To achieve the target sample size (N=20), a total of 95 individuals were invited to participate in the study (21% response rate). Participants included 10 females and 10 males, ranging in age from their mid-20s to late 70s. Of the 20 participants, nine had genetic testing related to inherited cardiomyopathy or arrhythmia syndromes, seven related to hereditary cancer risk, two related to a neurological indication (Cerebral Autosomal Dominant Arteriopathy with Subcortical Infarcts and Leukoencephalopathy [CADASIL]), and two related to other adult-onset conditions (retinitis pigmentosa and an autoinflammatory disorder) (**Fig. 1B**). All individuals underwent indication-based testing based on FH (n=3), PH (n=13), or both (n=4) (**Table 2**).

**Table 1.** Demographics and Genetic Testing Information.

| **Characteristic** | ***N*** |
| --- | --- |
| **Gender** |  |
| Female | 10 |
| Male | 10 |
| **Age** |  |
| <50 years | 6 |
| ≥50 years | 14 |
| **Age (range, years)** | 28 - 77 |
| **Time from reclassification to interview (range, months)\*** | 1 - 22 |

\*Time since reclassification was calculated by taking the difference between the time when reclassified report was generated and the time when participant agreed to the interview

**Table 2.** Testing indication and genes with reclassified result.

| Participant ID | Testing Indication | Gene(s) with reclassified variant | Reclassification Type | Time to Reclassification (months)* | Time from Reclassification to Interview (months) |
| --- | --- | --- | --- | --- | --- |
| P01 | Personal history of Brugada syndrome | *SCN5A* | VUS→LP | 42 | 6 |
| P02 | Personal history of HCM | *SPRED1* | VUS→B | 3 | 6 |
| P03 | Personal history of DCM | *DSP* | VUS→LP | 6 | 6 |
| P04 | Personal history of HCM | *MYBPC3* | VUS→LP | 5 | 7 |
| P05 | Personal history of HCM | *TTN* | VUS→LP | 24 | 6 |
| P06 | Personal history of HCM | *MYH7* | VUS→P | 24 | 8 |
| P07 | Personal and family history of HCM | *MYBPC3* | P→B | 50 | 2 |
| P08 | Family history of breast and pancreatic cancer | *CDH1* | LP→VUS | 1 | 4 |
| P09 | Personal history of pancreatic and ovarian cancer | *MSH6* | VUS→P | 10 | 5 |
| P10 | Personal history of breast cancer and family history of breast and prostate cancer | *MSH6* | VUS→B | 13 | 1 |
| P11 | Personal history of Retinitis Pigmentosa | *SNRNP200* | VUS→LB | 56 | 8 |
| P12 | Personal history of sebaceous adenoma | *MUTYH* | VUS→LP | 3 | 15 |
| P13 | Family history of breast cancer | *MLH1* | VUS→B | 2 | 15 |
| P14 | Personal history of DCM | *GAA* | VUS→P | 50 | 16 |
| P15 | Family history of CADASIL | *NOTCH3* | VUS→LP | 7 | 5 |
| P16 | Personal history suspicious for CADASIL | *NOTCH3* | VUS→LP | 41 | 6 |
| P17 | Personal history of pancreatic NET and uveal melanoma, and family history of lung cancer | *MSH2* | VUS→B | 9 | 3 |
| P18 | Family history of pancreatic cancer | *MEN1* | VUS→LB | 11 | 2 |
| P19 | Personal history of HCM | *MYH7* | VUS→P | 34 | 19 |
| P20 | Personal history of autoinflammatory symptoms without a definitive diagnosis | *STAT4* | VUS→LB | 2 | 22 |

\*Time to reclassification was calculated by taking the difference between the initial report date and the most relevant/recent amended report date. The total number of months was rounded to the nearest whole month. HCM - Hypertrophic Cardiomyopathy. DCM - Dilated Cardiomyopathy. CADASIL - Cerebral Autosomal Dominant Arteriopathy with Subcortical Infarcts and Leukoencephalopathy. NET - Neuroendocrine Tumor.

Interviews occurred between 1 and 22 months after the most recent reclassification was reported (median 6 months). Reclassification types included VUS upgraded to likely pathogenic or pathogenic (LP/P; n=11), VUS downgraded to likely benign or benign (LB/B; n=7), pathogenic downgraded to benign (n=1), and likely pathogenic downgraded to VUS (n=1) (**Fig. 1C**). The time from initial result disclosure to reclassification ranged from 1 to 56 months (median 11 months).

### Thematic analysis of patient responses to variant reclassifications

Thematic analysis resulted in three overarching themes characterizing patients’ responses and opinions to receiving a variant reclassification (**Box 1, Supplemental Table**).

### BOX 1

Theme 1: Effectiveness of the care team communicating variant reclassifications.

Theme 2: Varied perceptions of the value of genetic testing in light of a reclassification.

Theme 3: Reclassification as a manifestation of personalized medicine.

**Theme 1:** Effectiveness of the care team communicating variant reclassifications

More than half of the participants (n=11) expressed satisfaction with their providers’ communication of the reclassified results. Yet, a similar number (n=12) expressed that improvements are needed in how these results are communicated. Participants wanted less technical language in their explanations (n=2), continued access to the care team after the reclassification (n=2), proactive outreach and guidance from the care team after reclassification (n=5), and improved methods of communication of reclassified results (n=5). One participant who had a pathogenic to benign reclassification in *MYBPC3* expressed frustration that he received the results document prior to verbal disclosure:

> “*[…] Rather than just sending the doc, and then let me figure it out or guess what’s going on, it would have been good [if] somebody called and said, hey, this is what we are going to release and this high level, this is the reason results have changed, and then give me the paper doc, then I would have been okay*.”(P07; PH and FH of Hypertrophic cardiomyopathy (HCM); P-B in *MYBPC3*)

One participant, who had an initial VUS in *MSH2* gene that was later reclassified to benign, spoke about the need for results delivery that balances timeliness with providing appropriate context and support:

> *“I think the difficulty here, at least from my perspective, is getting the results in a medical scientific lab report format a long time before a talk with a doctor creates anxiety. But then also, hearing nothing until my next appointment with the doctor leaves me feeling kind of uncared for.”* (P17; PH of pancreatic neuroendocrine tumor (PNET) and uveal melanoma; VUS-B in *MSH2*)

Several (n=4) participants noted that they would have preferred a different method of communication depending on the type of reclassification. One participant who had an initial VUS in *MSH6* gene that was later reclassified to pathogenic noted:

> “*Well, looking back, I might have preferred having an in-person appointment, because that was pretty important news. […] immediately I thought that a positive result was positive, not negative. […] I have so many appointments on any given week that maybe I just said it’s fine to give me the results over the phone, not knowing what they were gonna tell me*.” (P09; PH of pancreatic and ovarian cancer; VUS-P in *MSH6*)

Another participant who had an initial VUS in *MSH6* gene that was later reclassified to benign also expressed:

> *“In general, I think if it’s bad news, it’s always better to have somebody deliver that to you in a phone call or some kind of personal contact. But if it’s something like this, where it was actually a confirmation of what I had been told at the time [*…*] finding it out via MyChart was fine.”* (P10; PH of breast cancer and FH of breast and prostate cancer; VUS-B in *MSH6*)

The direction of variant reclassification was also reflected in participants’ emotional responses to receiving the reclassified result. This was noticed when we looked at what the participants who had an initial VUS result reported when asked about their emotional response to the reclassified result. Participants whose VUS was downgraded to LB/B (n=7) described positive reactions, including feeling happy or reassured. In contrast, none of the participants whose variants were upgraded to LP/P (n=11) reported these emotions. One participant with a LB/B downgrade also reported a sense of remaining uncertainty after their reclassified result. Among those experiencing upgrades, participants described feeling overwhelmed, disappointed, confused, or angry. These negative emotions were not expressed by any of the participants with LB/B downgrades. Notably, feelings of validation and surprise were reported at similar frequencies across both groups (**Fig. 2**).

**Figure 2.**
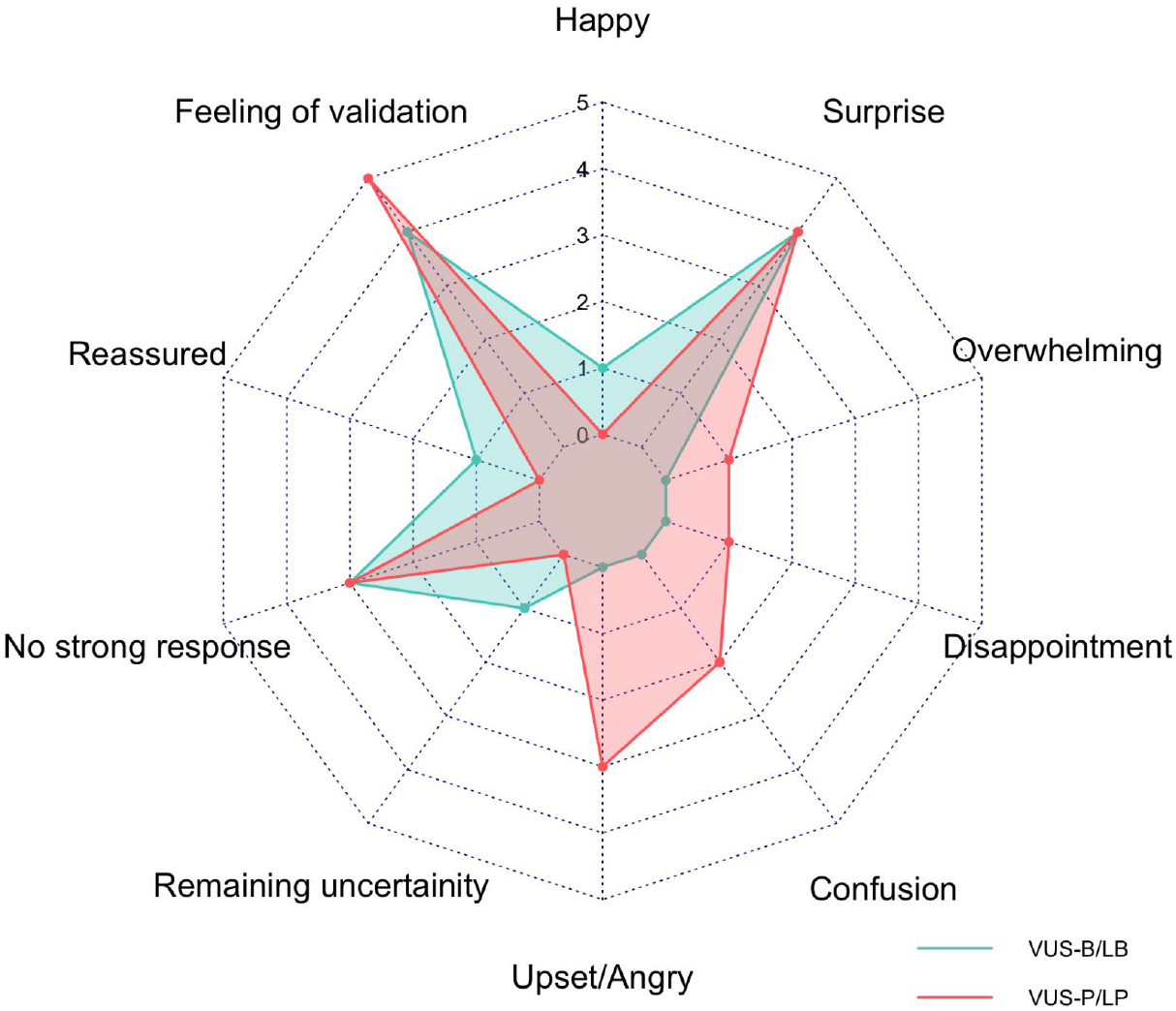
Emotional reaction to reclassified results for those who had an initial VUS. A radar chart showing different emotional reactions reported by participants who had an initial VUS separated by the direction of reclassification.

The time elapsed between the initial and reclassified results also influenced participants’ communication preferences. One participant who had an initial VUS in the *SNRNP200* gene that was reclassified to likely benign noted:

*“Initially, I got a text message and then I also got an email. But yeah, I thought the text message was kind of a scam [*…*] Because you know, it’s been several years since I thought about it, since I’ve talked to the doctors about it. [*…*] because it came out of nowhere and it’s an email, it felt too detached and impersonal to me.”* (P11; PH of Retinitis Pigmentosa; VUS-LB in *SNRNP200*)

This participant further explained that this experience created a barrier to reaching out to the genetics team for clarification about his result despite him not understanding the result fully:

> “*I was reading it [letter explaining reclassified result], but I didn’t really quite understand it too well. They gave me contact information to call to ask questions, but I never did [*…*] if I would have gotten a phone call instead, initially, even a voicemail, then I think that would have, that will have increased the chance that I would contact them about it [questions about the reclassified result].”* (P11; PH of Retinitis Pigmentosa; VUS-LB in *SNRNP200*)

Two other participants also expressed encountering barriers to following up with the genetics team after reclassification. One participant who had an initial VUS in the *MEN1* gene that was reclassified to likely benign noted:

> *“And honestly, I think that there’s an opportunity for me to reach back out to that genetic counselor. But I haven’t, and it’s just not been something that I’ve done, or it’s not easily attainable. I mean, I probably could go through my charts and look through a bunch of different things and try to figure it out, but it’s not readily available for me to reach out to that individual.”* (P18; FH of pancreatic cancer, VUS-LB in *MEN1*)

This same sentiment was expressed by another participant who had an initial VUS in the *TTN* gene that was reclassified to likely pathogenic:

> *“I remember seeing some up on MyChart and me going. Oh, that’s different. That’s new. I didn’t really expect anything else to come from it, and I also was like, Oh, I wonder if this like reopens the door to get my family tested. [But] because no one else reached out to me. I was like oh, I guess not.”* (P05; PH of HCM; VUS-LP in *TTN*)

### Theme 2: Varied perceptions of the value of genetic testing in light of a reclassification

Theme 2 was typified by participants commenting on how their experience with reclassification influenced their overall impression of the value of genetic testing. Most participants explicitly viewed genetic testing as worthwhile (n=15), and some held nuanced views, describing genetic testing as beneficial when factors such as confidentiality, cost, and educational background were carefully weighed (n=2).

For example, one participant was concerned about the implications of someone outside of the care team having access to her results:

> *“If medical insurance companies could have access to those results that could seriously impact who they were willing to insure, and what kind of premiums they’d have to pay, and that kind of stuff.”* (P14; PH of dilated cardiomyopathy; VUS-P in *GAA*)

Several participants (n=8) described how reclassification reinforced their appreciation for the ongoing scientific work behind genetic testing. One participant with a personal history of CADASIL and an initial VUS in *NOTCH3* that was reclassified to likely pathogenic noted:

> *“I appreciate that, as more research, as more technology catches up with things, that my genetic stuff is there, and I appreciate the technology and science.”* (P16; PH suspicious of CADASIL; VUS-LP in *NOTCH3*)

Another participant with an initial VUS in *MSH2 that was reclassified to benign noted:*

> *“I think, in general, it [the experience of going through genetic testing and receiving reclassification] helps inspire a greater level of confidence in the diagnostic and the, and the prognosis, and kind of the whole system, I think, just… it gives me greater confidence than not having that information. [*…*] The fact that that work [continued research] is happening makes me feel good.”* (P17; PH of PNET and uveal melanoma; VUS-B in *MSH2*)

Despite most participants appreciating the value of genetic testing, over one third of participants (n=7/20) expressed systemic and/or financial challenges in accessing initial genetic testing or genetics providers following reclassification. Three participants also expressed that they were surprised by the dynamic nature of genetic testing. One of these participants with a personal history of HCM and an initial pathogenic variant in *MYBPC3* had a positive view of genetic testing at the time of initial testing as the result explained the personal history of disease. However, the participant expressed a change in his opinion on the value and accuracy of genetic testing once the variant was downgraded to benign:

> *“Yeah, it is not accurate. It’s not accurate, and you cannot make decisions based on that. Maybe you can, but you cannot 100% rely on that.”* (P07; PH and FH of HCM; P-B in *MYBPC3*)

Another participant who had a downgrade from likely pathogenic to VUS in *CDH1* described that while he saw value in genetic testing, he still questioned the accuracy with which genetic test results are translated into clinical recommendations:

> *“So if anything could be taken away from this, do two genetic tests on people instead of one, because, you know it can be wrong, and even if it’s rarely wrong, it’s worth the second test.”* (P08; FH of breast cancer and pancreatic cancer; LP-VUS in *CDH1*)

### Theme 3: Reclassifications as a manifestation of personalized medicine

Theme 3 was typified by participants interpreting genetic test reclassifications as a reflection of personalized care. For example, 11 participants described feeling positively surprised that someone was keeping track of their genetic result, long after their initial testing. One participant with a personal history of HCM and an initial VUS in *TTN* that was reclassified to likely pathogenic noted:

> *“[Referring to regular doctor visits] You’re not really thought of until maybe 2 days before you go see a doctor [*..*] But here, it was like, oh, they are going back. They’re updating things. They are like revising the data and making changes to it. And I thought that was nice. In the sense of, I’m not just forgotten forever, because I tested a year or 2 years prior.”* (P05; PH of HCM; VUS-LP in *TTN*)

This sentiment of “I am not forgotten” was echoed by others who felt that the follow-up indicated that the medical system cared about their wellbeing. For example, one participant with an initial VUS in *SNRNP200* gene that was reclassified to likely benign noted:

> *“I was surprised, because I didn’t think they would continue looking into my sample […] and It kind of made me feel kind of hopeful, in a way, because it made me think that they were serious about researching my condition and trying further their understanding of it, and made me feel like they were trying to really help me and people like me.”* (P11; PH of Retinitis Pigmentosa; VUS-LB in *SNRNP200*)

A noteworthy point here was patients’ attitudes toward different types of reclassification results, particularly downgrades to LB/B. When asked, 92% (n=12/13) of participants strongly preferred being informed even when a VUS was downgraded. For example, a participant with an initial VUS in *STAT4* that was reclassified to likely benign noted:

> “*Of course [I would like to know]! Yeah, because it’s still part of my information. Yeah, that’s definitely valuable information*.” (P20; PH of autoinflammatory symptoms; VUS-LB in *STAT4*)

In another case where an initial VUS in *GAA* was upgraded to pathogenic, the participant stated that knowledge of a benign outcome would still be valuable in terms of reducing emotional burden:

> *“It will be one less marker to be concerned about. It kind of not necessarily dismisses it, but just alleviates some of the pressure.”* (P14; PH of dilated cardiomyopathy; VUS-P in *GAA*)

## Discussion

Through qualitative interviews with 20 patients who received a variant reclassification across a range of clinical indications and reclassification trajectories, this study examined patient experiences with variant reclassification as it currently occurs in routine clinical care. We identified three overarching themes. Theme 1 highlighted the need for improvement in how reclassified results are communicated, while Themes 2 and 3 underscored the broader relational and interpretive consequences of reclassification, including its impact on patients’ trust in genetic testing and biomedical research. Together, these findings suggest that variant reclassification is not only a technical reinterpretation of genomic data, but also a meaningful clinical and relational event that can shape how patients perceive genomic medicine.

In the 1999 ACMG policy statement on the “Duty to re-contact,” the authors presciently observed that “There are now many examples in the face of remarkable advances in human genetics in which genetic counseling given just a few years previously is now known to be incorrect”^21^. Variant reclassification represents one such consequence of the rapid progress in genomic medicine. A 2019 ACMG “Points to Consider” statement on “Patient re-contact after revision of genomic test results” offered guidance to health-care providers, clinical testing laboratories, and patients by framing re-contact as a “shared responsibility,”^10^ but left unresolved important practical questions, including how best to initiate re-contact, how reclassified results should be communicated in practice, and which types of variant reclassifications warrant patient re-contact. In our study, 17% of individuals (22/127) were not contacted to participate either because they had passed away or did not have documented communication about the reclassification. Of these, four individuals had a lab report with the reclassified result in their EHR but did not have any documented communication from the clinician, reflecting variable re-contact practices. This observation parallels recent findings documenting gaps in the dissemination of variant reclassifications, including 26 cases in which the testing laboratory updated ClinVar but the reclassification was never communicated to the patient^22^.

More than half of participants in our study expressed a desire for improved communication surrounding variant reclassifications. In current clinical practice, reclassified results are often issued directly by the testing laboratory, with updated reports released simultaneously to both patients (for example, through electronic portals) and ordering providers. However, this model compresses the timeframe in which genetics providers and ordering clinicians can review reclassified results, contextualize their implications, and re-engage patients in follow-up discussions. As noted in the original 1999 ACMG policy statement, under ordinary circumstances medical geneticists do not maintain ongoing relationships with patients and may face substantial challenges in re-engaging patients years after initial testing. Our findings suggest that these structural realities can translate into patient experiences of uncertainty, anxiety, or perceived lack of support when reclassified results are released without adequate contextualization.

Strategies for improving re-contact may be informed by participants’ varying responses to the direction and clinical implications of the reclassification. Reclassifications involving an upgrade from B/LB/VUS to P/LP, or a downgrade from P/LP, were associated with participants describing feelings of being overwhelmed, disappointed, confused, or angry. These emotional responses were not reported by participants whose variants were downgraded from VUS to B/LB, which are commonly electronically returned to patients^23^ – a mode of communication that has been shown to be well received by patients^18^. More broadly, the appropriate mode of communication may also depend on whether the reclassified result is consistent with what was discussed during pre-test counseling, clinical diagnosis, or family history, as unexpected results may warrant more direct, supported communication^17^. Notably, several participants described hesitation in initiating follow-up discussions with genetics providers, particularly when those providers were not part of their ongoing care team. These findings suggest that reliance on patient-initiated follow-up may be insufficient, especially for reclassifications with significant clinical or emotional implications.

Taken together, our results support consideration of tiered or stratified re-contact approaches based on the nature of the variant reclassification. For reclassifications with greater potential to alter clinical management or provoke distress, such as upgrades to P/LP or downgrades from P/LP classifications, automatic scheduling of a formal clinical encounter, in addition to initial notification by phone or electronic communication, may better align with patient needs. Ensuring timely follow-up is particularly important given contemporary practices of immediate electronic release of laboratory reports, which almost always precede provider-mediated interpretation. However, systemic changes to workflows and billing structures may be required to implement this.

Our findings also underscore the broader implications that variant reclassifications can have on patients’ trust in genetic testing and biomedical research. As discussed in Rashkin et al. 2022, variant reclassification can cause harm even in the absence of negligence, at the same time showing that scientific discovery is working. Harm from reclassification can take multiple forms, including medical harms such as unnecessary procedures performed or necessary ones forgone, as well as financial and emotional consequences. It can also extend beyond patients to providers, who may fear that reclassification will cause harm to the patients and be perceived as prior error on their part. In our study, the majority of participants interpreted reclassification as a manifestation of precision medicine, fostering trust in genomic medicine. However, a subset of participants experienced negative emotional responses following reclassification. One participant voiced losing trust in the accuracy of genetic testing entirely, referring to it as “not accurate”, perhaps understandably given the magnitude of their reclassification (P-B). While a downgrade reclassification from P/LP to B/LB may be individually uncommon, they are not negligible. Rashkin et al. estimate the probability of a P/LP being downgraded to range from approximately 1 in 110 to 1 in 1,000. These estimations show the importance of anticipating and addressing the consequences that accompany such downgrades. Several participants also expressed nuanced views, stating that genetic testing is valuable after a thorough understanding of its limitations and uncertainties. Some appeared surprised by the dynamic nature of genetic testing, suggesting that many patients are not adequately prepared for the possibility of reclassification **(Supplemental Table)**.

Effectively preparing patients for the possibility of reclassification requires intervention at multiple levels. Genetic providers routinely discuss the limitations and evolving nature of genetic testing during pre- and post-test counseling, yet our findings suggest that this information does not always translate into patient understanding or retention. This challenge is likely to intensify as pre-test discussions are increasingly initiated by non-genetics specialists with varying genetics expertise, and as reclassifications are subsequently triaged to genetics providers years later. The gap between what is communicated and what is internalized points to the need for solutions beyond the clinical encounter alone. Broader public education through school curricula or public health efforts to improve general genetic literacy is needed to build a foundation that allows patients to contextualize reclassification not as a failure of medicine or false positive, but as evidence that our understanding continues to grow. Additionally, the evolving landscape of genomic medicine underscores the need for defined recontact pathways at the institutional level to reduce reliance on individual clinicians, and to minimize the risk of undisclosed reclassifications as patients transition between providers or specialties

Notably, we show that participants do not uniformly interpret reclassification as a failure. Across Themes 2 and 3, multiple participants described reclassification as reinforcing their appreciation for the ongoing scientific work underlying genetic testing and as signaling that the medical system continued to care about their wellbeing. These perspectives highlight variant reclassification as a potential opportunity to strengthen trust and relationships between patients and biomedical research in ways that extend beyond the immediate clinical encounter. Importantly, participants emphasized that these benefits were not limited to variant upgrades, but also extended to VUS downgrades, with 92% (n=12/13) of participants who were asked strongly preferring to be informed when a VUS was downgraded. Although VUS are typically managed clinically in a manner similar to B/LB variants—and the clinical value of re-contact following such downgrades remains debated, leading some laboratories not to routinely notify patients or providers^11^—our findings are consistent with prior research suggesting that returning these results may carry broader psychosocial and relational benefits for patients^24,25^.

While our study recruited a balanced cohort across testing indications and gender, several limitations must be considered. Since the study sample was drawn from two academic medical centers and limited to English-speaking adults, these findings may not fully generalize to more diverse patient populations. This study was limited to adults, so it is unclear if these results will generalize to the pediatric setting. Additionally, while we captured a wide breadth of experiences across various reclassification types, we did not systematically collect data on ethnicity, socioeconomic background or health literacy, factors known to shape how patients navigate genetic testing^26–29^. Finally, while the qualitative analysis was grounded in rigorous methodology, it remains an interpretive process informed by the lens of the research team. To build on these findings, future research should examine variant reclassification within more diverse populations and across a broader range of clinical and social environments.

Overall, our study highlights concrete pathways for improving how patients are re-contacted following variant reclassification and emphasizes the importance of viewing reclassification not solely as a laboratory update, but as a consequential moment in patient care. Thoughtfully designed re-contact practices have the potential not only to reduce uncertainty and distress, but also to reinforce trust, transparency, and patient engagement in personalized genomic medicine.

## Supporting information

Consenting and Interview Guide

## Data Availability

The authors confirm that the data supporting the findings of this study are available within the article or in the supplemental materials provided.

## Author Contributions

Conceptualization: A.B.S., S.K., P.G., M.S.P.; Data Curation: P.G., M.S.P.; Formal Analysis: P.G., M.S.P., S.K., A.B.S.; Funding Acquisition: A.B.S., L.M.S., D.M.F ; Investigation: P.G., M.S.P.; Methodology: P.G., M.S.P., S.K., A.B.S.; Project Administration: P.G., M.S.P., M.H.-P., A.B.S.,; Resources: A.B.S., E.Y.K., M.H-P, P.G., M.S.P.; Software: P.G., M.S.P.; Supervision: A.B.S., S.K.; Visualization: P.G., M.S.P.; Validation: P.G., M.S.P., A.B.S., S.K., A.E.M., R.D.K.; Writing-original draft: P.G., M.S.P., S.K., A.B.S.; Writing-review & editing: P.G., M.S.P., E.Y.K., A.E.M., R.D.K., D.M.F., L.M.S., S.K., A.B.S.

## Ethics Declaration

The study (STUDY00022283) was conducted in accordance with the Declaration of Helsinki (1975) and in compliance with 45 Code of Federal Regulations (CFR) 56 for studies involving human participants. Waivers of consent and Health Insurance Portability and Accountability Act (HIPAA) were obtained from the University of Washington Biomedical Institutional Review Board. Individual-level data, including clinical data, were deidentified.

## Conflict of Interest

The authors declare no conflicts of interest.

## Acknowledgments

The authors thank all the participants who agreed to be interviewed for this study and Martha J. Horike-Pyne for her support in navigating IRB procedures. This work was presented at the UW Medical Genetics Clinic Conference and the 2025 UW Genomic Medicine Research Symposium.

## Funding

This study was supported by a National Institutes of Health grant (R01HG013025, A.B.S., L.M.S., A.E.M., and D.M.F..), and a Chan Zuckerberg Initiative Foundation grant (CZIF2024-010284, A.B.S, D.M.F. and L.M.S.). A.B.S. holds a Career Award for Medical Scientists from the Burroughs Wellcome Fund and is a Pew Biomedical Scholar. A.E.M. was supported by an Early Career Award from the Alex’s Lemonade Stand for Childhood Cancer and RUNX1 foundation (21-25037), and the Brotman Baty Institute Catalytic Collaborations Grant (CC28). P.G. was supported by the Career Ladder Education Program for Genetic Counselors grant from the Warren Alpert Foundation (WAF-CLEP-PD 10089501-01).

## Notes

### Competing Interest Statement

The authors have declared no competing interest.

### Author Declarations

This study was approved by the Institutional Review Board (IRB) at the University of Washington (STUDY00022283).

