## Supplementary material for "Not Forgotten: Patient Experiences with Genetic Variant Reclassifications": Consenting and Interview Guide

### BBI-CVD Qualitative Study Consenting Script and Interview Guide

#### Introduction and Consent

**When Patient contacts the study.** Hi, I'm XXX, a genetic counselor at the Uni of WA. Yes, you have reached the research study. Thank you for contacting us regarding this research. I can explain the study.to you. We are contacting people who get their care at (UWMC / FH) and who received a genetic test result that was recently updated. We are conducting a research project on patient's feelings about, and experiences in, receiving genetic risk reports that were changed with new information. Did you receive updated or re-classified genetic test results?
.

**When Patient told Provider that contact was OK.** Hi, Is this XXX? I'm XXX, a genetic counselor at the Uni of WA. Do you remember telling Dr. XXX that it would be OK to call you to ask if you want to learn more about this project?). Did you receive updated or re-classified genetic test results? Is this a good time to talk? We are contacting people who get their care at (UWMC / FH) and who received a genetic test result that was recently updated. We are conducting a research project on patient's feelings about, and experiences in, receiving genetic risk reports that were changed with new information.

**When Patient has no previous information on the research.** Hi, Is this XXX? I'm XXX, a genetic counselor at the Uni of WA. Is this an OK time to tell you about a new research project of interviews to gain understanding of patient's perspective on receiving updated or reclassified genetic test results? Did you receive updated or re-classified genetic test results? We are contacting people who get their care at (UWMC / FH) and who received a genetic test result that was recently updated. We are conducting a research project on patient's feelings about, and experiences in, receiving genetic risk reports that were changed with new information.

Is this a good time for you to proceed with the approximately 30 to 45-minute interview? y/n—schedule or proceed).Thank you for taking the time to speak with me today.

I'll first describe the research project to you, and if you want to participate, I'll need to get your verbal consent to talk with me as a research participant. You have no obligation to participate in the interview, and there are no consequences if you decide to decline.

This research study is funded by the NIH being conducted by the University of Washington School of Medicine. We are conducting these 30 to 45-minute interviews are part of a project focused on learning patient's feelings about genetic testing, and the whole experience of getting genetic test results. Specifically, we are interested in how patient's feel, or their emotions when they first receive the genetic test results and then, subsequently, learn that their those results have been updated and changed. This process of updating genetic results is called reclassification or re-interpretation what the genetic risk results, which can mean a change in risk of developing a future disorder or disease. We are only interviewing people who received an update or change in their genetic risk results that is different from what their first or initial test result indicated. We will not be talking about the medical importance of the change, only about your feelings in receiving a changed or updated report. Thus, our conversation will have no medical benefit to you, but may help us understand patient's perspectives. We will record our conversation to allow us to capture all that was said. We will transcribe the conversation, and then destroy the actual tape.

The interview questions will ask things like: 1) How did you learn of your first genetic result report,  and how did you feel after getting a later update result 2) Did you talk to anyone about your results, why or why not? 3) We'll ask if you have any suggestions for improvements in delivering genetic results.

- Is it clear what types of questions will be asked? Remember, you can refuse to answer any question or stop the interview at any time.

Are you interested in continuing and having an interview with me? I'll need to obtain your consent to continue and to record your consent and our conversation.

To make sure the research is thoroughly and accurately described (*Points to make).*

- You are a volunteer. You can decline to answer or discuss any topic. You can ask any question at any time.
- I will tell you when we start and stop recording
- None of your responses will be shared with your doctor or anyone else in your medical team.
- There is no payment to you for your time.
- Your responses will be taped, then the tape destroyed after creating a written transcription. We will keep your information private until the end of the research, stored without identifiers.

- **Do I have your permission to audio tape our conversation?** .

- Your risks for participation are primarily a potential loss of privacy for your opinions.
- There is also a risk in our conversation around genetic test results that might be upsetting for you. Remember, just say "pass" for any question you don't want to answer.
- Again, you can expect no benefits for your participation---the benefit is primarily to society, if learning patient's opinions and observations make for better delivery of genetic results.
- After the interview, I will send you an email with my contact information in case you have any questions or think of something you meant to say. Is this all clear and do you have any questions for me before we begin?

- **Do I have your permission and consent to start the Interview?**

  Would you like to think about it and answer me later? [*Proceed as appropriate), **Write down consenting date, time and interviewers initials).***

---

#### Interview

- **May I turn on the recorder now to record our conversation? [START RECORDING]**

This interview is on (date) at (time) with participant (name). He/She has consented into being interviewed. Is that correct (name) that you consent to participating in this research? (participant affirms).

- **<u>Questions related to Initial result:</u>** These questions all relate to you getting your first genetic test result that was categorized as a _______ (classification noted in the initial clinical report).

    - It looks like you had genetic testing performed in <month, year>. What do you remember about your initial genetic testing? [tell the participant the correct classification of the variant if what they remember is incorrect]

Thinking about when you were told, and how your felt, when you received your first genetic test result.

1. Tell me about how you were told and how you felt when you received the first genetic test result.
2. How did the doctor explain the “variant classification on initial report” and what it meant for your health?
    a. How well did the doctor explain the result? Did you feel like you understood everything? Why or why not?
    b. How would you like to have your doctor explain or discuss the result with you?
    c. Where did you look for other information about your result? What sources of information were most helpful?
        *Prompt: web searches, social media, advice from friends or family, documentaries, news, books/magazines, academic papers, etc.*
3. [If the initial classification was a VUS] At that time, what were your expectations for eventually receiving updated, more certain information about your genetic test results?

4. Have you made any changes to your behavior or lifestyle based on your updated results? What are they? Why did you make these changes?

5. Have you talked with your family about what your updated results mean for them? What have those conversations been like?

- **<u>Questions related to reclassified result</u>:** These questions all relate to you getting your updated genetic test result that was changed.

    1. Do you remember what your test result was recently updated or reclassified to? <classification noted in the updated clinical report – “positive/pathogenic”, “negative”, or “uncertain/VUS” >, (help as needed)

- **<u>Questions related to emotional response</u>**: These questions relate to how you felt about the changed results.

1. How did you feel when you received the updated result?
    a. What was your emotional reaction?

- ❖ **<u>Questions related to comprehension, communication, behavior:</u>** Let's talk a bit about how you learned about the updated result.

1. How did the doctor explain the updated result and what it meant for your health?
    a. How well did the doctor explain the updated result? Did you feel like you understood everything? Why or why not?
    b. How would you like to have your doctor explain or discuss the result with you?
    c. Where did you look for other information about your updated result?
        i. *Prompt: web searches, social media, advice from friends or family, documentaries, news, books/magazines, academic papers, etc.*

1. Have you made any changes to your behavior or lifestyle based on your updated results? What are they? Why did you make these changes?

2. Have you talked with your family about what your updated results mean for them? What have those conversations been like?

- ❖ **<u>Questions related to trust in Medical System</u>**: I'm also interested in understanding how receiving a result that was labeled uncertain significance, and later was updated with new information.

1. How did the updated information impact your view of genetic testing and the medical system overall?

2. In what ways has this updated experience influenced your views about the value of genetic testing?

    a. Probe: What would you tell a friend or family member who asked you whether or not to get genetic testing.

3. Did you feel like you had sufficient support from your doctor and/or team of doctors during the process of receiving an uncertain result that was then updated?

    a. What types of support were most helpful?

    b. In what areas would you have benefitted from more support?

4. If you could provide our team with one piece of advice to improve the experiences of patients who receive uncertain genetic test results, what would it be?

- ❖ **<u>Additional thoughts/questions</u>:** Is there anything else you'd like to add about your experience or any other thoughts that have come up during our conversation?

<u>[Closing]:</u>

**** I'm turning the recorder off now. [STOP RECORDING]**

I will follow up with an email with my contact information in case you have any questions in the future or anything to add.

Thanks again for speaking with me today, I really appreciate you taking the time. Have a great rest of your day!
